# Population level impact of a pulse oximetry remote monitoring programme on mortality and healthcare utilisation in the people with covid-19 in England: a national analysis using a stepped wedge design

**DOI:** 10.1101/2021.11.29.21266847

**Authors:** T Beaney, J Clarke, A Alboksmaty, K Flott, A Fowler, JR Benger, P Aylin, S Elkin, AL Neves, A Darzi

**Author notes:** Corresponding Author: Dr Thomas Beaney, Patient Safety Translational Research Centre, Institute of Global Health Innovation, Imperial College London, London, SW7 2AZ, United Kingdom.

## Abstract

**Objectives:** To identify the population level impact of a national pulse oximetry remote monitoring programme for covid-19 (COVID Oximetry @home; CO@h) in England on mortality and health service use.

**Design:** Retrospective cohort study using a stepped wedge pre- and post-implementation design.

**Setting:** All Clinical Commissioning Groups (CCGs) in England implementing a local CO@h programme.

**Participants:** 217,650 people with a positive covid-19 polymerase chain reaction test result and symptomatic, from 1^st^ October 2020 to 3^rd^ May 2021, aged ≥65 years or identified as clinically extremely vulnerable. Care home residents were excluded.

**Interventions:** A pre-intervention period before implementation of the CO@h programme in each CCG was compared to a post-intervention period after implementation.

**Main outcome measures:** Five outcome measures within 28 days of a positive covid-19 test: i) death from any cause; ii) any A&E attendance; iii) any emergency hospital admission; iv) critical care admission; and v) total length of hospital stay.

**Results:** Implementation of the programme was not associated with mortality or length of hospital stay. Implementation was associated with increased health service utilisation with a 12% increase in the odds of A&E attendance (95% CI: 6%-18%) and emergency hospital admission (95% CI: 5%-20%) and a 24% increase in the odds of critical care admission in those admitted (95% CI: 5%-47%). In a secondary analysis of CO@h sites with at least 10% or 20% of eligible people enrolled, there was no significant association with any outcome measure. However, uptake of the programme was low, with enrolment data received for only 5,527 (2.5%) of the eligible population.

**Conclusions:** At a population level, there was no association with mortality following implementation of the CO@h programme, and small increases in health service utilisation were observed. Low enrolment of eligible people may have diluted the effects of the programme at a population level.

## Background

Since the start of the covid-19 pandemic, asymptomatic (‘silent’) hypoxia has complicated the assessment and care of patients with covid-19.[1] Hypoxia has been shown to be an important predictor of mortality and the need for hospital admission in patients with covid-19, yet those patients with asymptomatic hypoxia may be unaware of dangerously low blood oxygen saturations.[2,3] Pulse oximetry allows patients and clinicians to regularly monitor a patient’s oxygen saturation and promptly initiate escalation of treatment should deterioration occur.[1] Health systems across the world introduced remote monitoring pathways, including the use of pulse oximetry, to support the care of patients with Covid-19 outside hospital.[4,5] In November 2020, NHS England and Improvement (NHSEI) introduced the COVID Oximetry @home (CO@h) programme, recommending that all Clinical Commissioning Groups (CCGs) in England provide services to monitor patients with a diagnosis of covid-19 at home using pulse oximetry.[6] This service builds upon local remote monitoring services provided by individual CCGs in earlier stages of the pandemic. CCGs are responsible for the establishment of services in their area, although CO@h services may be shared between CCGs, and more than one service may operate within a single CCG.[7]

Patients are eligible for enrolment into a CO@h programme if they have symptomatic covid-19 and are aged 65 years or older or are identified as clinically extremely vulnerable (CEV) based on concurrent medical conditions and/or treatments.[8] Additionally, clinical judgement can be applied to consider other individual risk factors, including pregnancy, learning disability, and socioeconomic deprivation. Those enrolled are encouraged to record three oximetry readings daily and advised to attend or call emergency services if the reading is 92% or less, or to contact primary care services for readings of 93-94%. The programme is intended to accept patients from primary care, NHS Test and Trace, ambulance services and accident and emergency departments, in contrast to ‘COVID Virtual Wards’ which aim to support discharge of covid-19 patients from hospital.[9]

The clinical effectiveness of the CO@h programme on mortality and secondary care utilisation remains unknown. The primary aim of this analysis is to identify the impact of the CO@h programme on mortality and use of healthcare services at a population level amongst people eligible for the programme. A secondary aim is to identify the impact of the programme in sites with a high uptake of the programme amongst the eligible population.

## Methods

This study used a retrospective cohort of people eligible for the CO@h programme, comparing outcomes at a CCG level using a stepped wedge pre- and post-implementation design. Eligibility was defined as the population resident in England, with a positive covid-19 Polymerase Chain Reaction (PCR) test result, who were symptomatic at the time of testing, from 1^st^ October 2020 to 3^rd^ May 2021. Of these individuals, a subset of people who would have remained eligible throughout the course of the programme, aged 65 years or older, or who were Clinically Extremely Vulnerable (CEV) was selected. Care home residents were excluded from the analysis, as previous work has suggested significantly higher mortality in this group.[10]

Five primary outcomes were selected, defined as occurring within 28 days of the date of a positive covid-19 test:

1. Death from any cause
2. One or more Accident and Emergency department (A&E) attendances
3. One or more emergency hospital admissions
4. One or more critical care admissions (of those admitted to hospital)
5. Total hospital length of stay in days, of those admitted who did not die within 28 days

### Data sources and processing

Covid-19 testing data was provided through the Public Health England Second Generation Surveillance System[11], which collates positive covid-19 test results conducted in laboratories across England. Data was available from 1^st^ October 2020 to 3^rd^ May 2021. This analysis used PCR tests, with symptoms documented at the time of testing.[12] Where more than one test was recorded, only the date of first test was used. Data on the number of patients enrolled (‘onboarded’) onto the CO@h programme were submitted from CO@h sites via NHS Digital’s Strategic Data Collection Service (SDCS).[13] Primary care data was sourced from the General Practice Extraction Service (GPES) Data for Pandemic Planning and Research (GDPPR).[14] Hospital Episode Statistics (HES) data[15] and the Emergency Care Data Set[16] provided data on hospital admissions and A&E attendances up to 31^st^ May 2021. Data on registration of deaths was sourced from the Office for National Statistics (ONS), with data available up to 5^th^ July 2021. Datasets were linked using a deidentified NHS patient ID.

The study population were assigned to a CCG based on a person’s CCG of residence when the test was performed. Patient demographic data, including age, sex, ethnicity, lower layer super output area (LSOA) of residence were derived from GDPPR, or, if missing, derived from HES or ECDS. LSOA was linked to measures of deprivation based on the Index of Multiple Deprivation (IMD) 2019.[17] Data on care home residence, CEV status, body mass index (BMI) and smoking status were available from GDPPR only. Information on the following chronic conditions were included, derived from GDPPR: hypertension, chronic cardiac disease, chronic kidney disease, chronic respiratory disease, dementia, diabetes, chronic neurological disease (including epilepsy), learning disability, malignancy/immunosuppression, severe mental illness, peripheral vascular disease and stroke/transient ischaemic attack (TIA). For demographics and chronic conditions, the most recent codes prior to the date of a positive covid-19 test were used to exclude those which may have resulted from covid-19 infection. If no data were available prior to the date of a positive test for age, sex and ethnicity only, then the earliest data following the positive test was used. Full details of the datasets and cleaning approach are provided in Appendix A, with a link to the Systematized Nomenclature of Medicine Clinical Terms (SNOMED-CT) codes for each condition in appendix A.

### Statistical analysis

A pre- and post-implementation period were defined for each CO@h site, with implementation start dates for each site provided by NHS England @home. A stepped wedge design was used. All eligible people in each CCG before and after implementation of the CO@h programme were allocated to the control group and intervention group, respectively. Two-level hierarchical regression models were run for each outcome, incorporating random intercepts for CCG. Logistic regression was used for the four binary endpoints and negative binomial regression models were used for the single continuous outcome (length of stay).

Total length of stay was capped at 28 days where a patient was discharged after the 28-day window, and analyses of length of stay excluded patients who died within the 28-day time window.

To account for possible changes in the baseline risk of each outcome over time, the primary models for each outcome incorporated fixed effects for the month of positive covid-19 test. To account for potential differences in the at-risk population before and after implementation, the primary models adjusted patient-level risk factors. Final covariates in each model included age category (years), sex, ethnicity, IMD score, BMI category, month of covid-19 test, CEV status, and clinical co-morbidities. Intra-class correlation coefficients were calculated for each model.

To test the sensitivity of results to time-varying effects and differences in patient risk factors, a series of sensitivity analyses were run:

1. A naïve model, not accounting for time or patient-level covariates
2. A time-adjusted model, without adjusting for patient-level covariates
3. A time-adjusted model, incorporating a random interaction between time and CCG
4. A time-adjusted model incorporating a random interaction between time and CCG, and adjusting for patient-level covariates

A secondary analysis was performed on the subset of sites with a higher proportion of eligible people enrolled. Two thresholds were defined *a priori*, at 10% or more (accounting for sixteen CCGs and 9.4% of the eligible population) and 20% more (accounting for five CCGs and 2.4% of the eligible population) across the whole study period.

Analyses were conducted in the Big Data and Analytics Unit Secure Environment (BDAU SE), Imperial College. Python v3.9.5 and Pandas v1.2.3 were used in data manipulation. Regression models were conducted in Stata v17.0, using the *melogit* and *menbreg* commands.

### Patient and public involvement

Patients or the public were not involved in the design, conduct or reporting of our research.

## Results

A total of 1,714,182 people resident in 106 CCGs in England had a positive PCR covid-19 test between 1 October 2020 and 3rd May 2021, with documented symptoms at the time of the test. Eligibility for the CO@h programme was defined as a person being at least 65 years of age or CEV, resulting in an eligible population of 223,429 (13.0%). Of these, 5,779 (2.6%) were living in a care home at the time of their positive test and were excluded from the analysis. A total of 217,650 people were included in the final analysis (Figure 1).

**Figure 1:**
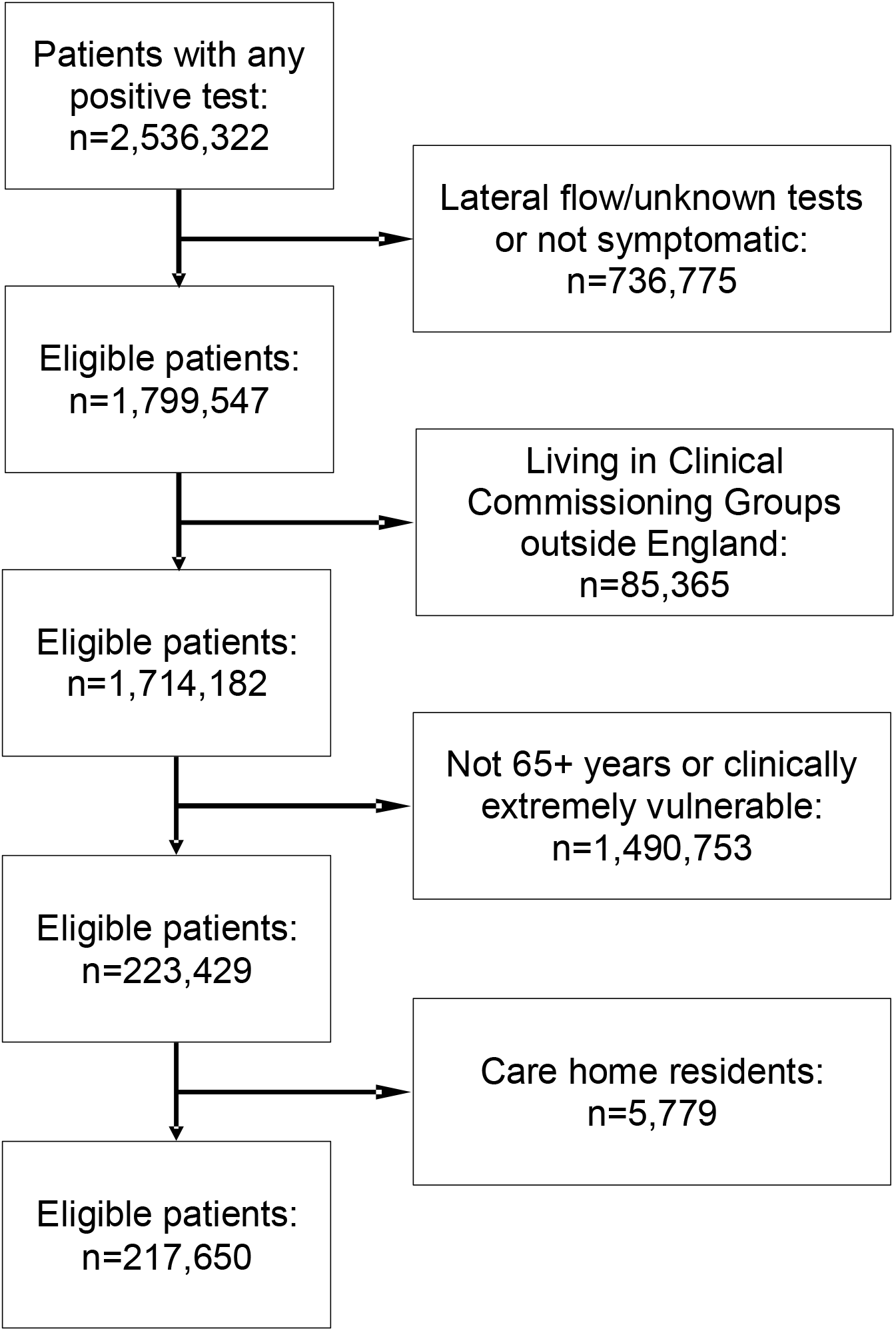
Flow diagram of eligibility criteria for the evaluation of the CO@h programme.

Most participants were women (54.4%) and of White ethnic background (72.5%), with 125,102 (57.5%) in the five most deprived deciles. More of the population were obese (39.4%) than overweight (33.6%) or had a healthy weight (20.8%). Just over half (54.9%) were never smokers. Hypertension (40.6%), diabetes (31.4%; type 1 and type 2) and chronic respiratory disease (29.3%) were the most common co-morbidities. A total of 5,616 (2.6%) of the study population died within 28 days of a positive covid-19 test, 19.9% attended A&E at least once, 12.2% were admitted at least once, and of those admitted, 16.1% required critical care. There were significant differences in distributions of most of the predictor and outcome variables in the eligible population before and after implementation in each site (Table 1).

**Table 1:** Characteristics of people eligible for the CO@h programme from 1 October 2020 to 3rd May 2021, before and after implementation at each site.

|  | Pre-implementation |  | Post-implementation |  | p-value for difference |
| --- | --- | --- | --- | --- | --- |
|  | Number | Percentage | Number | Percentage |  |
| Age category (years) and Clinically Extremely Vulnerable status |  |  |  |  |  |
| 18-49 and CEV | 15364 | 20.5% | 33138 | 23.2% | p<0.001 |
| 50-64 and CEV | 10319 | 13.8% | 21219 | 14.9% |  |
| 65-79 and not CEV | 36401 | 48.6% | 64181 | 45.0% |  |
| 65-79 and CEV | 5726 | 7.6% | 10010 | 7.0% |  |
| 80+ and not CEV | 4875 | 6.5% | 9270 | 6.5% |  |
| 80+ and CEV | 2215 | 3.0% | 4932 | 3.5% |  |
| Sex |  |  |  |  |  |
| Female | 39701 | 53.0% | 78610 | 55.1% | p<0.001 |
| Male | 33896 | 45.3% | 61759 | 43.3% |  |
| Missing | 1303 | 1.7% | 2381 | 1.7% |  |
| Ethnicity |  |  |  |  |  |
| White | 58112 | 77.6% | 99703 | 69.8% | p<0.001 |
| Asian/Asian British | 10725 | 14.3% | 25757 | 18.0% |  |
| Black/African/Caribbean/Black British | 1631 | 2.2% | 6755 | 4.7% |  |
| Mixed/Multiple ethnic groups | 685 | 0.9% | 1865 | 1.3% |  |
| Other ethnic group | 1076 | 1.4% | 3472 | 2.4% |  |
| Missing | 2671 | 3.6% | 5198 | 3.6% |  |
| Index of Multiple Deprivation (IMD) decile |  |  |  |  |  |
| 1 (most deprived) | 11697 | 15.6% | 16548 | 11.6% | p<0.001 |
| 2 | 9207 | 12.3% | 18218 | 12.8% |  |
| 3 | 8038 | 10.7% | 17379 | 12.2% |  |
| 4 | 7233 | 9.7% | 15678 | 11.0% |  |
| 5 | 6884 | 9.2% | 14220 | 10.0% |  |
| 6 | 6510 | 8.7% | 13565 | 9.5% |  |
| 7 | 6691 | 8.9% | 12722 | 8.9% |  |
| 8 | 6839 | 9.1% | 12191 | 8.5% |  |
| 9 | 6380 | 8.5% | 11855 | 8.3% |  |
| 10 (least deprived) | 5409 | 7.2% | 10337 | 7.2% |  |
| Missing | 12 | 0.0% | 37 | 0.0% |  |
| Body Mass Index |  |  |  |  |  |
| Underweight | 724 | 1.0% | 1646 | 1.2% | p<0.001 |
| Healthy weight | 15181 | 20.3% | 29999 | 21.0% |  |
| Overweight | 25648 | 34.2% | 47591 | 33.3% |  |
| Obese | 29770 | 39.7% | 56064 | 39.3% |  |
| Missing | 3577 | 4.8% | 7450 | 5.2% |  |
| Smoking status |  |  |  |  |  |
| Never smoker | 39901 | 53.3% | 79530 | 55.7% | p<0.001 |
| Ex-smoker | 24770 | 33.1% | 41668 | 29.2% |  |
| Current smoker | 8862 | 11.8% | 18852 | 13.2% |  |
| Missing | 1367 | 1.8% | 2700 | 1.9% |  |
| Co-morbidities |  |  |  |  |  |
| Hypertension | 30548 | 40.8% | 57810 | 40.5% | 0.194 |
| Chronic cardiac disease | 12482 | 16.7% | 22818 | 16.0% | p<0.001 |
| Chronic kidney disease | 1070 | 1.4% | 2276 | 1.6% | 0.003 |
| Chronic respiratory disease | 22514 | 30.1% | 41276 | 28.9% | p<0.001 |
| Dementia | 790 | 1.1% | 1973 | 1.4% | p<0.001 |
| Diabetes | 21558 | 28.8% | 46886 | 32.8% | p<0.001 |
| Chronic neurological disease (including epilepsy) | 3004 | 4.0% | 6326 | 4.4% | p<0.001 |
| Learning disability | 459 | 0.6% | 1037 | 0.7% | 0.002 |
| Malignancy or immunosuppression | 15553 | 20.8% | 29204 | 20.5% | 0.092 |
| Severe mental illness | 1358 | 1.8% | 3066 | 2.1% | p<0.001 |
| Peripheral vascular disease | 1381 | 1.8% | 2264 | 1.6% | p<0.001 |
| Stroke or TIA | 3680 | 4.9% | 7154 | 5.0% | 0.316 |
| Deaths within 28 days | 1476 | 2.0% | 4140 | 2.9% | p<0.001 |
| Patients with at least one ED attendance within 28 days | 9965 | 13.3% | 24285 | 17.0% | p<0.001 |
| Patients with at least one emergency admission within 28 days | 7539 | 10.1% | 18990 | 13.3% | p<0.001 |
| Critical care use of those admitted | 1248 | 1.7% | 3027 | 2.1% | 0.220 |
| **Total** | **74900** | **34.4%** | **142750** | **65.6%** |  |

Data was received via submissions from CO@h sites for 5,527 patients onboarded to the programme, giving an overall uptake rate based on submitted data of 2.5%. There was considerable variation in uptake across CCGs, ranging from 0.0% to 33.0%, with a median of 2.2% (Figure 2). The earliest date a site became operational was 20^th^ November 2020, with all sites operational from 10^th^ January 2021.

**Figure 2:**
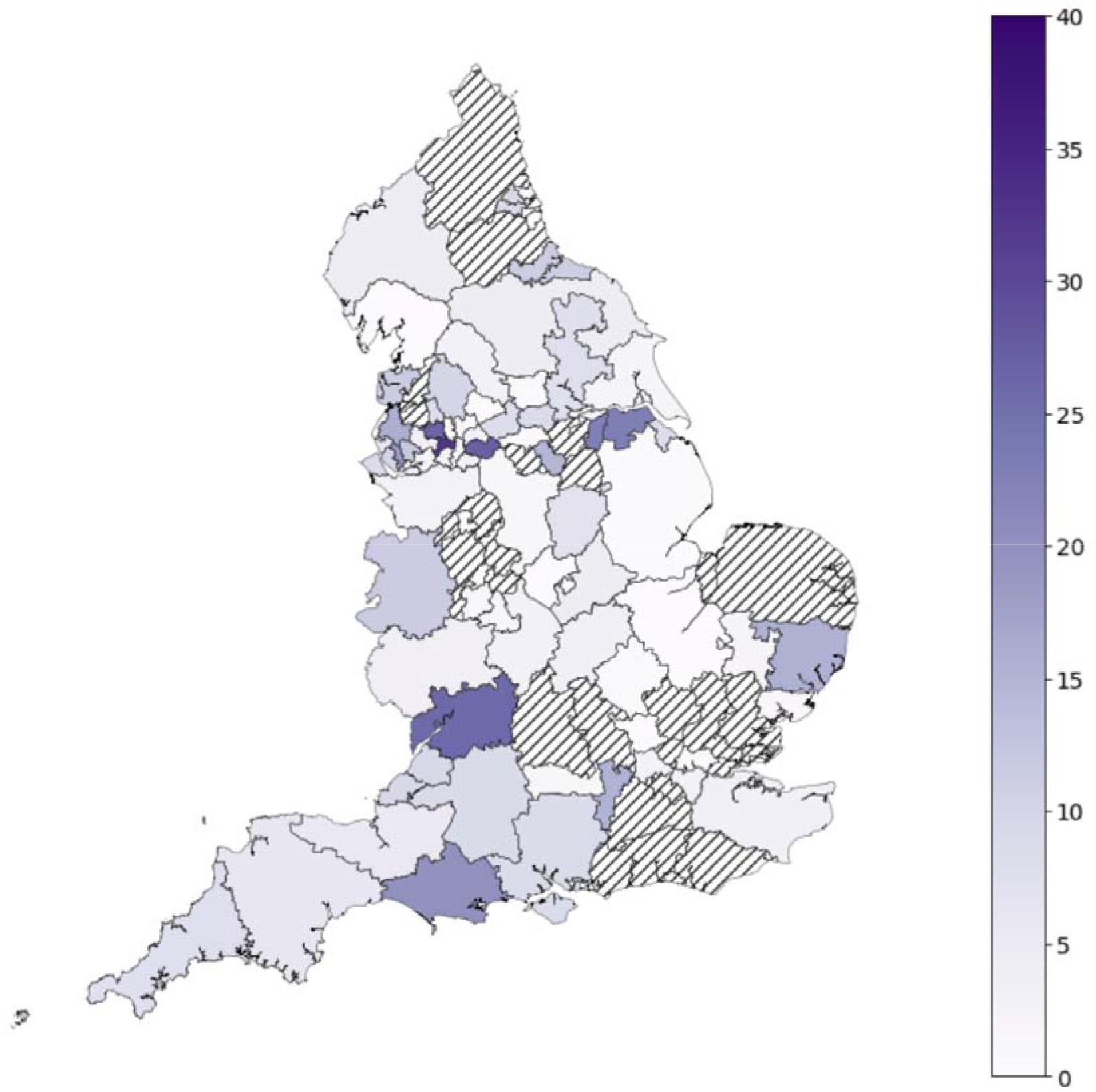
Percentage of the eligible population enrolled onto the CO@h programme in each CCG from date of implementation based on submissions from sites. *Hashed areas represent CCGs with no patient onboarding data submitted

Mixed effects logistic regression was run separately for each outcome, with CCG of residence as a random intercept. Table 2 shows the results for the primary analysis for each outcome, adjusted for month of covid-19 test and patient-level covariates. There was no significant difference in the adjusted odds of 28-day mortality following implementation of the CO@h programme (aOR=1.06, p=0.405). There was evidence of a small increase in both A&E attendances (aOR=1.12, p<0.001) and emergency hospital admissions (aOR=1.12, p<0.001) within 28 days. Of those patients admitted to hospital after implementation there was a 24% increase in the adjusted odds of requiring critical care (aOR 1.24, p=0.012). There was no significant difference in the length of stay of those admitted (p=0.588).

**Table 2:** Effect measures for the CO@h programme, from mixed effects regression models, adjusted for month of test and patient-level covariates.

| Outcome | Adjusted odds ratio | Standard error | p-value | 95% confidence interval |  | Denominator |
| --- | --- | --- | --- | --- | --- | --- |
|  |  |  |  | Lower | Upper |  |
| 28-day mortality | 1.06 | 0.072 | 0.405 | 0.93 | 1.21 | 203,218 |
| Any A&E attendance within 28 days | 1.12 | 0.033 | <0.001 | 1.06 | 1.18 | 203,218 |
| Any hospital admission within 28 days | 1.12 | 0.037 | <0.001 | 1.05 | 1.20 | 203,218 |
| Critical care use of those admitted | 1.24 | 0.107 | 0.012 | 1.05 | 1.47 | 24,895 |
|  | Adjusted IRR* | Standard error | p-value | Lower | Upper | Denominator |
| Length of stay of those admitted (in days) | 1.02 | 0.029 | 0.588 | 0.96 | 1.07 | 20,794 |

\*IRR = Incidence Rate Ratio from negative binomial regression

### Sensitivity analyses

Sensitivity analyses comparing alternative model specifications are given in the Appendix (Tables A1-5). Naïve models, unadjusted for time, showed significant increases in 28-day mortality, ED attendance and admissions associated with the programme. In contrast, in naïve models, the programme was weakly associated with lower odds of critical care admission. Little meaningful difference was seen in the estimates between models unadjusted or adjusted for patient-level covariates, or with the addition of random time by CCG interactions. The intraclass coefficients for both CCG and CCG by time interactions for mortality, ED attendance and hospital admission models were all <1%, suggesting minimal variation between CCGs that might be accounted for by time-varying CCG level factors.

### Secondary analysis of high uptake CCGs

Two secondary analyses using the same stepped wedge designed were performed for sixteen CCGs with 10% or more uptake (Table 4), and for five CCGs with 20% or more uptake (Table 5). In the 10% uptake group, there was a 9% lower odds or mortality, 10% higher odds of A&E attendance and 23% higher odds of critical care admission after implementation, but all effects were statistically non-significant. There was evidence of 27% higher odds of admission (p=0.046). In the 20% uptake CCGs, effect sizes were larger, but none were statistically significant.

**Table 3:** Effect measures for CO@h sites with 10% or more uptake, from mixed effects logistic/negative binomial regression models, adjusted for month of test and patient-level factors.

| Outcome | Adjusted odds ratio | Standard error | p-value | 95% confidence interval |  | Denominator |
| --- | --- | --- | --- | --- | --- | --- |
|  |  |  |  | Lower | Upper |  |
| 28-day mortality | 0.91 | 0.225 | 0.715 | 0.56 | 1.48 | 19,724 |
| Any A&E attendance within 28 days | 1.10 | 0.120 | 0.369 | 0.89 | 1.37 | 19,724 |
| Any hospital admission within 28 days | 1.27 | 0.151 | 0.046 | 1.00 | 1.60 | 19,724 |
| Critical care use of those admitted | 1.23 | 0.373 | 0.496 | 0.68 | 2.23 | 2,608 |
|  | Adjusted IRR* | Standard error | p-value | Lower | Upper | Denominator |
| Length of stay (days) of those admitted | 1.13 | 0.112 | 0.213 | 0.93 | 1.37 | 2,171 |

\*IRR = Incidence Rate Ratio

**Table 4:** Effect measures for CO@h sites with 20% or more uptake, from mixed effects logistic/negative binomial regression models, adjusted for month of test and patient-level factors.

| Outcome | Adjusted odds ratio | Standard error | p-value | 95% confidence interval |  | Denominator |
| --- | --- | --- | --- | --- | --- | --- |
|  |  |  |  | Lower | Upper |  |
| 28-day mortality | 0.89 | 0.397 | 0.798 | 0.37 | 2.13 | 4,807† |
| Any A&E attendance within 28 days | 1.27 | 0.248 | 0.216 | 0.87 | 1.86 | 4,887 |
| Any hospital admission within 28 days | 1.44 | 0.317 | 0.102 | 0.93 | 2.21 | 4,887 |
| Critical care use of those admitted | 1.64 | 0.954 | 0.393 | 0.53 | 5.13 | 652 |
|  | Adjusted IRR* | Standard error | p-value | Lower | Upper | Denominator |
| Length of stay (days) of those admitted | 1.27 | 0.259 | 0.246 | 0.85 | 1.89 | 552 |

\*IRR = Incidence Rate Ratio. †80 observations excluded in April/May as no deaths occurring

## Discussion

At a population level, there was no impact on mortality after implementation of the CO@h programme. Implementation was associated with statistically significant increases in A&E attendances and emergency hospital admissions, with a 12% increase in the odds of both A&E attendance or admission within 28 days of a positive test. There was some evidence of an increase in the odds of receiving critical care in patients admitted after implementation of the CO@h programme. In a secondary analysis of sites with at least 10% or at least 20% of eligible people onboarded, there was a trend towards lower odds of death and higher odds of A&E attendance, hospital admission and critical care after implementation, but effects were non-significant.

Increases in A&E attendances, emergency hospital admissions and critical care use following implementation of CO@h may reflect early recognition of silent hypoxia in covid-19. The magnitude of increase in A&E attendances was similar to the magnitude of increase in hospital admissions, suggesting that implementation did not cause a large increase in A&E attendances not requiring admission. However, early intervention might be expected to decrease length of stay and mortality, which was not found here. Despite this, the finding of no significant difference in mortality after implementation suggests that the CO@h pathway is safe and does not cause harm. Uptake of the programme was lower than expected, with data received from only 2.5% of the eligible population. This may be due to incomplete data submissions or may reflect genuinely low uptake of the programme. It could also reflect additional patient selection criteria used by sites, for example, if sites were focussing on patients with higher acuity disease. If uptake were only 2.5%, then there is likely to be a dilutional effect when evaluating results at a population level and we cannot infer a causal relationship at an individual level.

Remote monitoring technologies have been widely used in the management chronic diseases, but with mixed evidence of their clinical effectiveness[18] and there is limited evidence for their use in covid-19 with which to compare our findings (Alboksmaty, A *et al*. Effectiveness and safety of pulse oximetry in remote home monitoring of COVID-19 patients: a systematic review. Unpublished, under review at The Lancet Digital Health). A previous pilot study of four NHS covid-19 pulse oximetry programmes in England indicated the pathway was safe, but did not include a control group to compare outcomes.[19] A study in the US of patients with covid-19 referred to a remote patient monitoring pathway after discharge from hospital found lower odds of emergency department or hospital reattendance in those enrolled, but did not assess mortality.[20] Furthermore, the characteristics of patients discharged from hospital with covid-19 may be different to people in the community eligible for CO@h, who are likely to be at an earlier stage of disease.

In a separate study of CO@h programme by the same authors using a retrospective matched cohort design, patients with covid-19 enrolled after assessment in A&E were found to have lower odds of mortality and critical care use, and higher odds of subsequent A&E use and admission, compared with matched controls.[21] Collectively, results from the two studies suggest that although there was no impact on mortality at a population level, there is evidence for the effectiveness of the programme at an individual level (albeit within a narrower eligibility cohort of patients assessed in A&E), but indicate that the programme could not be scaled up to provide a benefit to all eligible people nationally. Crucially, neither of the studies indicate a concern over the clinical safety of the programme or that it causes harm. There is a need for further research into the barriers and facilitators to implementation of the programme and the obstacles to wider adoption which may aid policy-makers and commissioners in implementing remote monitoring programmes in the future. There is also a need to understand the equity of access to CO@h, and whether low enrolment nationally is reflected in lower numbers onboarded in certain sociodemographic groups, and to evaluate user experience and the cost implications of the programme.

### Strengths and limitations

A strength of this study is the availability of comprehensive data on covid-19 testing, defining a robust denominator population. Through linkage to primary and secondary care records, we identified the eligible population and adjusted for underlying risk factors differing between the population pre- and post-implementation. Eligibility for the programme was not absolute, with NHS England recommending an extended age threshold of 50 years and over from February 2021. Our analysis focussed on a narrower subgroup of people aged 65 or over or CEV, who would have remained eligible over the full study period, but as a result, findings might not be generalisable to all those included in the programme.

The stepped wedge design was chosen in part as it is robust to biases in decisions regarding onboarding of patients (such as systematic onboarding of higher or lower risk patients), which would impact individual level study designs. The effect estimates are also not impacted by lack of submission of patient-level programme data on patients onboarded, which may not be complete. However, incompleteness of received data does impact on our ability to judge the degree to which the study was adequately powered to detect a difference in mortality should one exist. The primary analysis adjusted for patient covariates, for which data were incomplete. However, the impact on the estimates is unlikely to be significant, as sensitivity analyses adjusted for patient-level covariates produced very similar estimates to the primary analysis.

Results may also be confounded by events external to the programme. A separate study using the same data sources showed significant increases in the case hospitalisation and fatality risk from covid-19 from October 2020 to January 2021 at the time CO@h sites were becoming operational.[22] These trends may relate to winter effects on mortality, new treatments, hospital pressures and spread of the alpha variant which became the dominant covid-19 strain in England during December 2020.[23,24] Despite the stepped wedge analysis incorporating time as an adjusting covariate, there may be residual confounding between sites becoming operational, and the underlying increase in mortality rates, particularly given the low numbers onboarded. Incorporation of additional CCG-level and hospital-level covariates was considered but estimates of the residual variation explained by clustering at the CCG-level (intra-class correlation coefficient) were less than 1%, suggesting these would have limited impact. The primary analysis considered time-varying CCG level effects in additional sensitivity analyses for each outcome, with almost identical effect measures compared to the primary analysis, suggesting little impact of time-varying differences between CCGs.

Across England, CO@h sites implemented different types of model, run by different sectors of the healthcare system, and with different recommendations for the frequency of monitoring.[4] National population effect estimates as presented here may therefore mask variation in the effectiveness between programmes and so not be representative of individual sites. Our analysis incorporated outcomes measured up to 28 days from the date of a positive covid-19 test, and different effects may be seen over longer time periods. Longer term outcomes, such as ‘Long Covid’ might also be affected by early recognition of low oxygen saturations but were outside of the scope of this study.

## Conclusion

Implementation of the CO@h programme across England had no impact on mortality at a population level and was associated with small increases in A&E attendances, hospitalisations and critical care use in people with covid-19 aged 65 years or over or CEV, which may indicate early recognition of hypoxia and escalation. Lower than expected enrolment of eligible people may have diluted the effects of the programme at a population level. There is a need for further research into the uptake and effectiveness of remote monitoring programmes for covid-19.

## Data Availability

The patient level data used in this study are not publicly available but are available to applicants meeting certain criteria through application of a Data Access Request Service (DARS) and approval from the Independent Group Advising on the Release of Data.

## Acknowledgements

The authors would like to thank Hutan Ashrafian, Gianluca Fontana, Saira Ghafur, Melanie Leis and Mahsa Mazidi for their input and support. Data management was provided by the Big Data and Analytical Unit (BDAU) at the Institute of Global Health Innovation (IGHI), Imperial College London. This work was funded by NHS England and supported by the National Institute for Health Research (NIHR) Imperial Patient Safety Translation Research Centre. Infrastructure support was provided by the NIHR Imperial Biomedical Research Centre (BRC). JC acknowledges support from the Wellcome Trust (215938/Z/19/Z).

## Ethics approval

The work was conducted as a national service evaluation of the CO@h programme, approved by Imperial College Health Trust on 3^rd^ December 2020. Data access was approved by the Independent Group Advising on the Release of Data (IGARD; DARS-NIC-421524-R0Y3P) on 15^th^ April 2021.

