## Supplementary appendix A for "Population level impact of a pulse oximetry remote monitoring programme on mortality and healthcare utilisation in the people with covid-19 in England: a national analysis using a stepped wedge design"

Corresponding Author:

Dr Thomas Beaney

**Data cleaning**

Note: the described datasets were used for several distinct analyses by the study team, with data cleaning rules the same or similar between studies. For this reason, some of the text in the appendices may be identical to that of other published articles by the study authors on the same data source.

*Covid-19 testing data*

Testing data was provided through the Public Health England Second Generation Surveillance System (SGSS). This dataset captures routine laboratory data on infectious diseases for England, including Covid-19, with all diagnostic laboratories required to notify positive test results within 24 hours.^1^ Data included 3,251,225 tests performed from 1^st^ October 2020 to 30^th^ June 2021, inclusive.

Provided data included test date and result date. 99% of results were reported within 5 days of the test, and 6,544 (0.2%) were reported more than 7 days from date of test. Where the result data occurred before the test date. In 1,603 cases, the result date was recorded as prior to the test date. In these instances, where the difference between the testing date and reporting date was 7 days or less, the test date and reporting date were swapped. In the 191 instances where reporting date was more than 7 days before the testing date, the test was excluded.

For this analysis, only tests performed up to 3^rd^ May 2021 were included (after swapping test and result dates where applicable), given that secondary care data was available up until the end of May 2021. Of the 2,928,802 positive Covid-19 tests, 2,352,390 (80.3%) were from Pillar 2 testing, 561,852 (19.2%) from Pillar 1, and 14,560 (0.5%) from Pillar 4.^2^ Test type was recorded for Pillar 2 tests only, and of these, 2,250,288 (95.7%) were Polymerase Chain Reaction (PCR) tests, 102,102 (4.3%) were lateral flow tests. Of PCR tests, 1,859,053 (82.6%) were documented as symptomatic at the time of test.

2,536,322 individuals had one or more positive Covid-19 tests. This analysis used only those with a positive PCR tests from pillar 2 testing, where symptoms were documented at the time of test, resulting in a total population of 1,799,547 people.

*COVID Oximetry @home (CO@h) programme data*

Data on patients enrolled (‘onboarded’) onto the CO@h programme were submitted directly from CO@h sites via NHS Digital’s Strategic Data Collection Service.^3^ Data included a deidentified NHS patient ID of the patient onboarded, along with the date of onboarding to and offboarding from the programme.

*COVID Oximetry @home (CO@h) sites*

Start dates at which each CO@h site became operational were provided by the NHS England @home programme at a Clinical Commissioning Group (CCG) level. Start dates were available for all CCGs in England, except for NHS commissioning hubs, which were excluded (National Commissioning Hub 1 (code 13Q), London Commissioning Hub (code 13R) and National Commissioning Hub 2 (code 15L)). The population was assigned to the CCG recorded in the SGSS dataset at the time of their test, and those outside of England were excluded. A final eligible cohort of 1,714,182 individuals with a PCR positive, and symptomatic at the time of test, were included

*Primary care data*

Primary care data came from the General Practice Extraction Service (GPES) Data for Pandemic Planning and Research (GDPPR).^4^ Data included month and year of birth, sex, ethnicity, Lower Layer Super Output Area (LSOA) of residence, a marker for Clinically Extremely Vulnerable (CEV) status, and a marker for residence in a care home. LSOA was used to link to 2019 deciles of Index of Multiple Deprivation (IMD).^5^ Age was calculated from date of positive Covid-19 test, assuming a birthdate on the 15^th^ day of the month.

Entries include a date to which each journal item applies, and a date on which the journal item was recorded. The former was used in priority, but where missing, was replaced with the journal item recording date. For LSOA, CEV status and care home residence, only entries occurring up to the date of positive Covid-19 test were included. For month and year of birth, sex and ethnicity, if no entry were included prior to the date of Covid-19 test, then the earliest recorded entry after the test was included.

*Secondary care data*

Data on hospital admissions came from the Hospital Episode Statistics (HES) data set up to 31st May 2021, linked to Office for National Statistics (ONS) data on death registrations up to 5th July 2021.^6^ Entries were excluded where missing admission dates, provider Trust code, or patient deidentified ID.

Where multiple admission episodes were recorded within a spell, a single spell start and end date were created. Non-emergency hospital admissions were excluded from analyses. A binary indicator was created for any admission within 28 days of positive Covid-19 test. A second indicator was created for death (of any cause) within 28 days of positive Covid-19 test. Critical care admissions were defined as an admission episode for any level 2 or level 3 care within the defined spell.

Data on accident and emergency (A&E) attendances came from the Emergency Care Data Set (ECDS).^7^ The A&E attendance date, departure date and admission date (if subsequently admitted) are included. In cases where attendance date was recorded as being after the departure date, the attendance date was set to the departure date, if departure date was equal to the admission date. Otherwise, attendance date was assumed to be correct.

Where multiple A&E attendances were recorded on the same day, a single attendance was kept for each patient, prioritising in turn:

1. Any attendance associated with an admission
2. Earliest time of attendance
3. Earliest time of departure

A binary indicator was created for one or more A&E attendances within 28 days of a positive Covid-19 test.

Where age was missing from GDPPR, it was derived from month and year of birth in HES, or if also missing in HES, derived from month and year of birth in ECDS, using the same approach as for GDPPR. Where LSOA was missing from GDPPR, it was derived from HES/ECDS. CCG was derived first from testing data, and if missing, from CO@h programme data, followed by, in order of use, GDPPR, HES or ECDS.

*Co-morbidities*

SNOMED codes were included in the GDPPR dataset pertaining to specific SNOMED code cluster reference sets provided by NHS Digital.^4^ 6,485 unique codes were identified from GDPPR. Codes were reviewed manually by authors TB and JC and removed if not relevant or assigned to the minimal number of relevant code clusters.

SNOMED reference clusters were aggregated into hierarchies of similar conditions. Codes in each higher-order cluster were then reviewed to ensure groupings of relevant codes and twelve relevant chronic disease categories were selected: hypertension, chronic cardiac disease, chronic kidney disease, chronic respiratory disease, dementia, diabetes, chronic neurological disease (including epilepsy), learning disability, malignancy/immunosuppression, severe mental illness, peripheral vascular disease and stroke/transient ischaemic attack (TIA). Categories for chronic respiratory disease, diabetes, epilepsy, malignancy/immunosuppression and severe mental illness included relevant medication codes. Broad diagnostic categories of diagnoses were chosen, as certain medications were not diagnostic of more granular diagnostic categories (for example, use of a long-acting bronchodilator/inhaled corticosteroid in both COPD and asthma) A full list of codes within each diagnostic category are available in Appendix B.

For each patient in GDPPR, all relevant diagnostic codes prior to the study index date (date of positive Covid-19 test) were considered diagnostic. In cases where the latest SNOMED code indicated resolution of a condition (e.g. ‘Atrial fibrillation resolved (finding)’), then the diagnosis was excluded for that patient. SNOMED codes relating to drug codes were only included up to 2 years prior to the index date.

*BMI categorisation*

SNOMED codes for BMI were either diagnostic categories (eg ‘Body mass index 30+ - obesity (finding)’ or value codes (e.g. ‘Body mass index (observable entity)’). Values were extracted and BMI was categorised according to the standard World Health Organisation classification of underweight (<18.5 kg/m^2^), healthy weight (18.5-24.9 kg/m^2^), overweight (25.0-29.9 kg/m^2^) and obese (≥30.0 kg/m^2^). Value codes outside of the range 5.0-100.0 kg/m2 were excluded. SNOMED codes which spanned more than one category (e.g. ‘Increased body mass index (finding)’) and child BMI categories were also excluded.

*Smoking categorisation*

Smoking status was categorised into ‘never-smoker’, ‘ex-smoker’ and ‘current smoker’ according to the latest SNOMED code prior to and including the index date. For any patient where the latest SNOMED code indicated ‘never-smoker’, but a prior record indicated active smoking, then the patient was re-categorised as ‘ex-smoker.

**Table A1: Effect sizes for the CO@h programme on 28-day mortality from mixed effects logistic regression models under different model specifications (n=217,650)**

| **Model** | **Odds ratio** | **Standard error** | **p-value** | **95% confidence interval** | | **Denominator*** |
| --- | --- | --- | --- | --- | --- | --- |
|  |  |  |  | **Lower** | **Upper** |  |
| Naïve | 1.53 | 0.05 | <0.001 | 1.44 | 1.63 | 217,650 |
| *Time adjusted models* | | | | | | |
| Simple | 1.07 | 0.068 | 0.259 | 0.95 | 1.22 | 217,650 |
| Full adjustment for patient factors | 1.06 | 0.072 | 0.405 | 0.93 | 1.21 | 203,218 |
| *Random time by CCG interaction* | | | | | | |
| Simple | 1.07 | 0.069 | 0.291 | 0.95 | 1.22 | 217,650 |
| Full adjustment for patient factors | 1.06 | 0.072 | 0.407 | 0.93 | 1.21 | 203,218 |

*Denominators lower in adjusted models due to missingness in patient-level covariates

**Table A2: Effect sizes for the CO@h programme on any A&E attendance within 28 days from mixed effects logistic regression models under different model specifications (n=217,650)**

| **Model** | **Odds ratio** | **Standard error** | **p-value** | **95% confidence interval** | | **Denominator*** |
| --- | --- | --- | --- | --- | --- | --- |
|  |  |  |  | **Lower** | **Upper** |  |
| Naïve | 1.31 | 0.018 | <0.001 | 1.27 | 1.34 | 217,650 |
| *Time adjusted models* | | | | | | |
| Simple | 1.13 | 0.032 | <0.001 | 1.07 | 1.19 | 217,650 |
| Full adjustment for patient factors | 1.12 | 0.033 | <0.001 | 1.06 | 1.18 | 203,218 |
| *Random time by CCG interaction* | | | | | | |
| Simple | 1.12 | 0.033 | <0.001 | 1.06 | 1.19 | 217,650 |
| Full adjustment for patient factors | 1.11 | 0.035 | 0.001 | 1.04 | 1.18 | 203,218 |

*Denominators lower in adjusted models due to missingness in patient-level covariates

**Table A3: Effect sizes for the CO@h programme on any hospital admission within 28 days from mixed effects logistic regression models under different model specifications (n=217,650)**

| **Model** | **Odds ratio** | **Standard error** | **p-value** | **95% confidence interval** | | **Denominator*** |
| --- | --- | --- | --- | --- | --- | --- |
|  |  |  |  | **Lower** | **Upper** |  |
| Naïve | 1.39 | 0.021 | <0.001 | 1.35 | 1.43 | 217,650 |
| *Time adjusted models* | | | | | | |
| Simple | 1.13 | 0.035 | <0.001 | 1.07 | 1.20 | 217,650 |
| Full adjustment for patient factors | 1.12 | 0.037 | <0.001 | 1.05 | 1.20 | 203,218 |
| *Random time by CCG interaction* | | | | | | |
| Simple | 1.13 | 0.036 | <0.001 | 1.06 | 1.20 | 217,650 |
| Full adjustment for patient factors | 1.12 | 0.038 | 0.001 | 1.05 | 1.20 | 203,218 |

*Denominators lower in adjusted models due to missingness in patient-level covariates

**Table A4: Effect sizes for the CO@h programme on any critical care admission of those admitted from mixed effects logistic regression models under different model specifications (n=26,529)**

| **Model** | **Odds ratio** | **Standard error** | **p-value** | **95% confidence interval** | | **Denominator*** |
| --- | --- | --- | --- | --- | --- | --- |
|  |  |  |  | **Lower** | **Upper** |  |
| Naïve | 0.92 | 0.036 | 0.04 | 0.85 | 1.00 | 26,529 |
| *Time adjusted models* | | | | | | |
| Simple | 1.15 | 0.093 | 0.087 | 0.98 | 1.35 | 26,529 |
| Full adjustment for patient factors | 1.24 | 0.107 | 0.012 | 1.05 | 1.47 | 24,895 |
| *Random time by CCG interaction* | | | | | | |
| Simple | 1.13 | 0.094 | 0.148 | 0.96 | 1.33 | 26,529 |
| Full adjustment for patient factors | 1.23 | 0.109 | 0.019 | 1.04 | 1.46 | 24,895 |

*Denominators lower in adjusted models due to missingness in patient-level covariates

**Table A5: Effect sizes for the CO@h programme on length of stay of those admitted from mixed effects negative binomial regression models under different model specifications (n=26,529)**

| **Model** | **Incidence rate ratio** | **Standard error** | **p-value** | **95% confidence interval** | | **Denominator*** |
| --- | --- | --- | --- | --- | --- | --- |
|  |  |  |  | **Lower** | **Upper** |  |
| Naïve | 1.04 | 0.015 | 0.004 | 1.01 | 1.07 | 22,079 |
| *Time adjusted models* | | | | | | |
| Simple | 1.02 | 0.029 | 0.495 | 0.96 | 1.08 | 22,079 |
| Full adjustment for patient factors | 1.02 | 0.029 | 0.588 | 0.96 | 1.07 | 20,794 |
| *Random time by CCG interaction* | | | | | | |
| Simple | 1.02 | 0.029 | 0.455 | 0.97 | 1.08 | 22,079 |
| Full adjustment for patient factors | 1.02 | 0.029 | 0.588 | 0.96 | 1.07 | 20,794 |

*Denominators lower in adjusted models due to missingness in patient-level covariates

6.	Digital, N. Hospital Episode Statistics (HES). https://digital.nhs.uk/data-and-information/data-tools-and-services/data-services/hospital-episode-statistics.

7.	NHS Digital. Emergency Care Data Set (ECDS). https://digital.nhs.uk/data-and-information/data-collections-and-data-sets/data-sets/emergency-care-data-set-ecds.
